# Outward and inward protections of different mask designs for different respiratory activities

**DOI:** 10.1101/2021.04.07.21255097

**Authors:** Xue Qi Koh, Anqi Sng, Jing Yee Chee, Anton Sadovoy, Ping Luo, Dan Daniel

## Abstract

We evaluate the outward and inward protections of different mask types (N95, surgical and two cloth mask designs) taking into account the imperfect fit on the wearer. To this end, we built a manikin to simulate exhaling, coughing and inhaling of aerosol droplets 0.3–5.0 *µ*m in diameters. The outward and inward protections depend on many factors, including the droplet size, the mask fit and the presence of a filter layer. Here, we show that cloth and surgical masks with a non-woven filter layer can achieve a combined outward and inward protections between 50% and 90%. Removing the filter layer greatly reduces the protection efficiency to below 20% for the smallest droplet size. While a well-fitted N95 masks offer protection efficiency close to 100%, a poorly fitted N95 mask with gaps offers less protection than a well-fitted surgical/cloth mask. We also found that double masking—the wearing of cloth mask on top of a surgical mask—is only effective at reducing outward droplet emissions when coughing, while offering no additional protection when exhaling/inhaling as compared to a single cloth/surgical mask. The results of our work can inform the implementation of mask mandates to minimize airborne transmissions of coronavirus disease of 2019 (COVID-19).

**Practical implications:**

1. A single cloth/surgical mask <u>with non-woven filter layer</u> offers significant protection against airborne transmissions.
2. Filtering facepiece masks such as N95 respirators are unlikely to achieve the advertised high protection for the general public due to poor mask fit.
3. Double-masking offers little to no additional protection over a single cloth/surgical mask.

## INTRODUCTION

There is a growing consensus that universal masking is an important tool to minimize airborne transmissions of coronavirus disease of 2019 (COVID-19) [1, 2]. Many countries now require the wearing of masks in public places. However, the implementation of mask mandates differ widely between jurisdictions: while some countries require their residents to only wear some form of face coverings including homemade cloth masks [3], others (notably France, Germany and Austria) have mandated the use of medical grade surgical masks and even filtering facepiece FFP2 masks [4].

While in theory FFP2 masks (and the equivalent N95 masks) can filter out more than 94% of 0.6 *µ*m particles [5], in practice the protections they provide are likely to be significantly lower for the general public, because minor differences in facial features, such as the presence of facial hair, can adversely affect the mask fit and hence performance [6]; healthcare workers are typically professionally trained and fitted for the right filtering facepiece with specialized instruments not available to the public. It is therefore unclear if there are additional benefits in mandating FFP2/N95 masks over high-quality surgical/cloth masks.

More recently, the Centers for Disease Control and Prevention (CDC) has advocated the wearing of cloth mask over surgical mask or double-masking to improve the mask fit. Brooks *et al*. (2021) found that double-masking can block close to 90% of simulated cough aerosol droplets, significantly more than the 60% blocking efficiency for a single cloth/surgical mask [7]. However, a recent computer simulation using the Japan’s Fugaku supercomputer did not support this conclusion [8].

In this paper, we would like to empirically examine the protection efficiencies of different masks and resolve some of the conflicting reports outlined above. To this end, we built a manikin to simulate exhaling, coughing and inhaling of aerosol droplets 0.3–5.0 *µ*m in diameters. We measured how well different mask types—surgical, cloth and N95 masks—as well as double-masking are able to block aerosol from the source (outward protection) and to minimize aerosol exposure to the receiver (inward protection) while taking into account imperfect mask fit. This is different from conventional particle filtration efficiency tests which look at how well the mask material is able to block droplets assuming a perfect mask fit with no gaps [9, 10].

While similar manikin set-ups have been reported by other groups [11–13], our approach differs in several important ways. Rather than generating a constant airflow, we chose to mimic respiratory activities more realistically by generating flow rates in bursts of between 0.3 and 3.0 s. In this way, we were able to evaluate the effects of flow rates on mask protections, which was not done in previous work. This is crucial because the flow rates generated by a cough/sneeze is significantly higher than exhalation cycle of human breath [14]. As we will show here, the effectiveness of mask protections depends on the exact respiratory activities.

## MATERIALS AND METHODS

### Aerosol generator

Aerosol typically refers to droplets with diameters less than 5 *µ*m and are thought to be responsible for airborne transmission of diseases, since they can stay suspended in air for a long period of time [15, 16]. Aerosol droplets generated during respiratory activities (such as talking and coughing) and in healthcare settings (such as intubation) are thought to vary between 0.1 and 10 *µ*m [16–18]. Here, we generate polydisperse aerosol droplets spanning this range from 2 wt% NaCl aqueous solution using an ultrasonic humidifier (home appliance) at room temperature of 22°C and 58–65% humidity.

The diameters of the droplets *D* generated vary between 0.3–5.0 *µ*m, as measured using MET ONE HHPC+ optical particle counter (See histogram in Fig. 1). Note that droplets below 0.3 *µ*m in size cannot be detected by the instrument. NaCl salt is added to mimic the solid content in respiratory fluid droplets and prevent the droplets from evaporating completely [19].

**FIG. 1.**
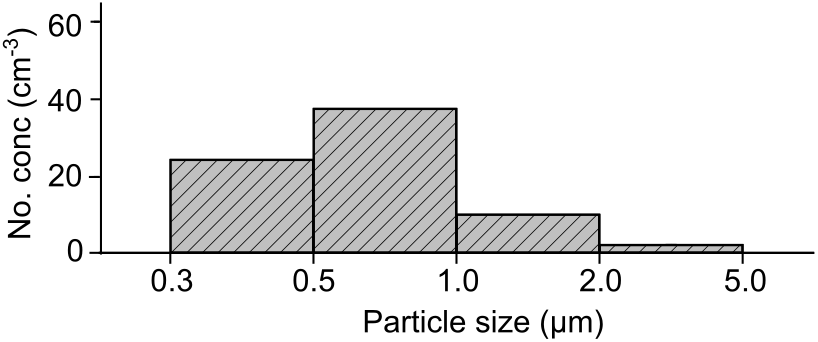
Distribution of aerosol droplets generated using an ultrasonic humidifier.

The total number of droplets generated can be tuned by adjusting the operation time of the humidifier. Millions of droplets can be produced within seconds.

### Simulating different respiratory activities

We connect a mechanical resuscitator or an Ambu bag to a life-sized manikin head with a 3 cm-sized orifice at the mouth region. Previously, we have used a similar set-up to quantify the outward protection of a barrier enclosure against airborne transmission [20].

To simulate exhalation and inhalation, we compress and re-inflate the Ambu bag fully by hand within Δ*t* = 3 s. The volume *V* expelled and drawn in with each compression and re-inflation cycle is determined to be 0.75 ± 0.05 L, generating a mean flow rate *Q* = *V/*Δ*t* = 15 L min^−1^ (International Organization for Standardization [ISO] standard for a female performing light work [7, 21]) and typical speed *U* ∼ 30 cm s^−1^. *V* is determined by releasing the expelled air into a jar filled with water and measuring the volume of water displaced. A metronome is used to keep the operator in time. We did not find significant differences in *V* for different human operators.

To simulate coughing, the Ambu bag was compressed by hand as quickly as possible. Experimentally, we found that *V* is expelled within Δ*t* = 0.30 ± 0.03 s to generate *Q* = 140 L min^−1^ or the equivalent *U* ∼ 3 m s^−1^, close to reported physiological parameters of human coughs [22]. Δ*t* was determined by recording the sounds of simulated coughs and measuring the time duration when the sound amplitudes are above background noise (Fig. 2). Previously, the sound of human coughs has been used to analyze cough duration and amplitudes [23].

**FIG. 2.**
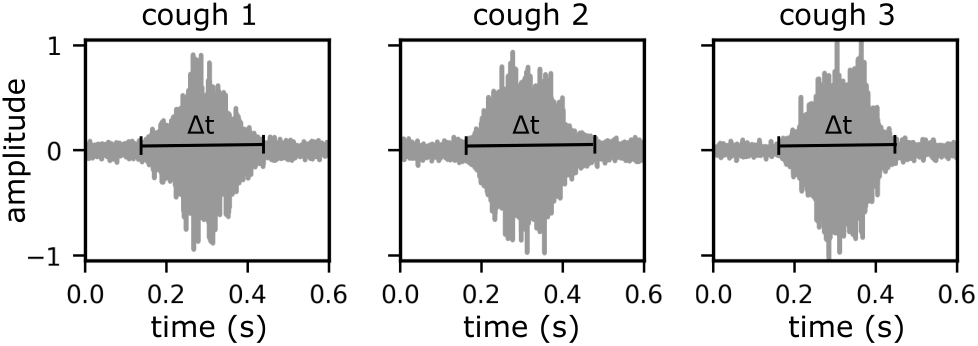
Sounds recorded during simulated coughs.

### Measuring outward and inward protections

The manikin head is placed inside an acrylic chamber with dimensions *w* × *l* × *h* = 0.6 0.5 0.6 m^3^ and volume *V*_1_ = 180 L (chamber 1 in Fig. 3). An Ambu bag connects the manikin in chamber 1 to a smaller chamber with dimensions *w* × *l* × *h* = 0.4 × 0.5 × 0.4 m^3^ and volume *V*_2_ = 80 L (chamber 2 in Fig. 3). Air can flow from one chamber to the other with the flow direction controlled by a one-way valve inside the Ambu bag.

**FIG. 3.**
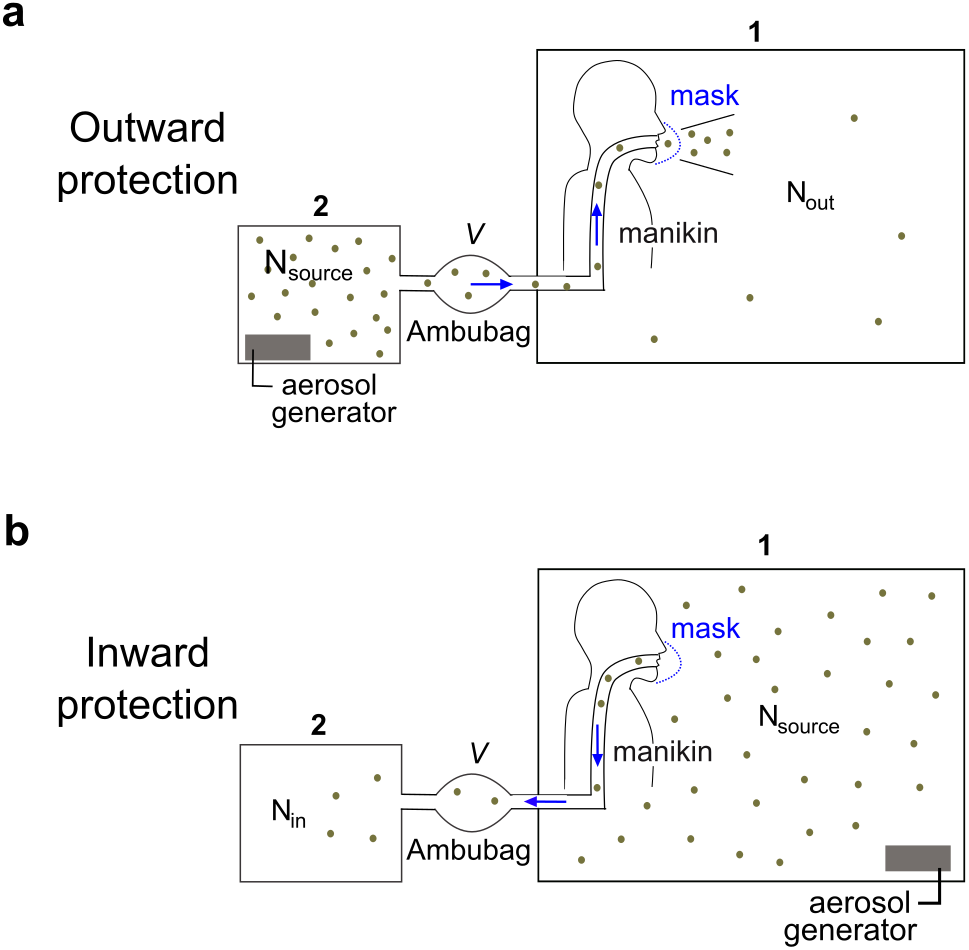
Experimental setup to measure (a) outward and (b) inward protections by different mask designs.

To measure the outward protection of masks *P*_out_, we first generate aerosols of different concentrations *ρ*_2_(*D*) inside chamber 2 (Fig. 3a). We then simulate four exhalation or cough cycles to transfer a portion of the aerosol *N*_source_ = 4*V ρ*_2_ from chamber 2 (the source chamber) to chamber 1. *P*_out_ can then be determined by comparing *N*_source_ with the number of droplets reaching chamber 1 and not filtered out by the mask *N*_out_ = *V*_1_*ρ*_1_, since

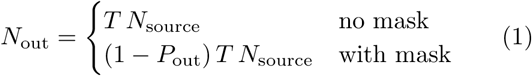

where *ρ*_1_ is the droplet concentrations in chamber 1 and *T* is the transfer rate of the droplets between the two chambers with no mask. *T* < 1 accounts for droplets that are deposited along the connecting pipes and the Ambu bag during aerosol transfer. To maximize *T*, we minimize the pipe lengths and avoid any bends in the pipes.

Since *N*_out_ ≤ *N*_source_, *ρ*_1_ ≤ 4*V/V*_1_ *ρ*_2_ = 0.02 *ρ*_2_, i.e. *ρ*_1_ << *ρ*_2_. Hence, we used two different optical particle sensor models (MET ONE HHPC+ and Sensirion SPS30) with suitable *ρ* range to accurately measure *ρ*_1_ and *ρ*_2_. The specifications of the two models are summarized in Table I. The optical particle sensors have four bin sizes which allow us to measure *P*_out_ for four different droplet size ranges: 0.4 ± 0.1, 0.75 ± 0.25, 1.5 ± 0.5 *µ*m and 3.5 ± 1.5 *µ*m. Note that the bin widths are slightly different for bins 3 and 4 for the two sensors, which we ignore in our analysis for simplicity. Throughout the experiment, small fans were used to distribute to the aerosol uniformly inside the two chambers.

**TABLE I.**
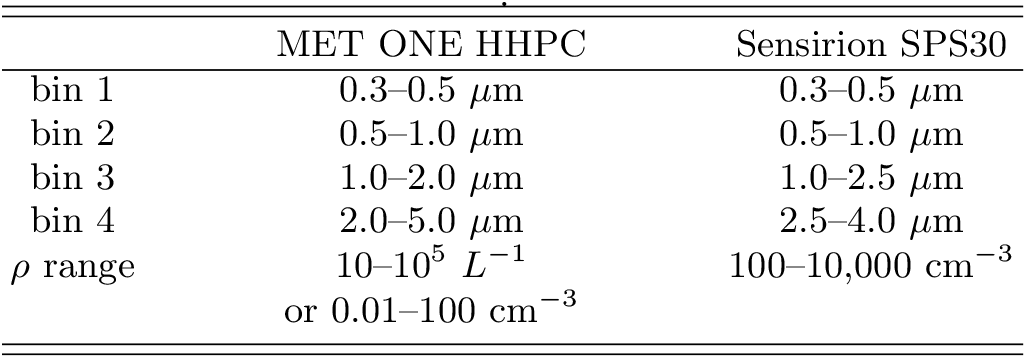
Specifications of the optical particle sensors used

A similar setup was used to measure the inward protection of masks *P*_in_, except the aerosol is now generated in chamber 1 and the direction of the flow is from chamber 1 to 2 (Fig. 3b). After four inhalation cycles, the protection efficiency is determined by comparing *N*_source_ = 4*V ρ*_1_ and the number of droplets reaching chamber 2 unfiltered *N*_in_ = *V*_2_*ρ*_2_, where *ρ*_1_ and *ρ*_2_ are measured using Sensirion SPS30 and MET ONE HHPC+ sensors, respectively. As before, we expect

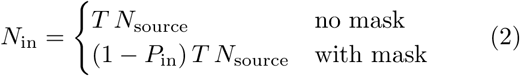

By varying the initial number of droplets from the source *N*_source_, we can obtain the slopes of *N*_out, in_ against *N*_source_. From equations 1 and 2, we deduce that

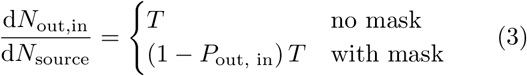

The outward and inward protections can then obtained through the relation

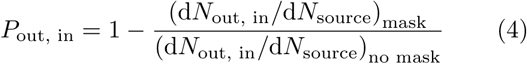

Experimental uncertainties in *P*_out, in_ depends on the uncertainties in the two slope values, i.e.

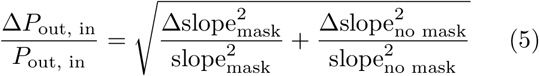

### Mask fit

In our experiments, the cloth/surgical mask is securely fastened using the ear loops to cover both the nose and mouth of the manikin snugly. The nose bridge is also adjusted to minimize gaps above the nose area. It is not possible however to completely eliminate gaps for cloth and surgical masks.

Healthcare workers typically undergo professional mask fitting to eliminate gaps around an N95 mask. To obtain a similar level of mask fit, we use a silicone sealant around the mask perimeter after securing it on the manikin. To simulate the lower level of protection for N95 masks worn by the general public, we mould the mask snugly onto the manikin but without applying the silicone sealant.

### Background particle levels

The number density of background particles *ρ* (*D*=0.5– 5.0 *µ*m) in our lab is about 1000 *L*^−1^. To minimize background *ρ*, we used a commercially available air ionizer from Zero2.5 Biotech, Singapore. Turning on the ionizer for 1 min before the start of each experiment reduces *ρ* < 10 *L*^−1^ insider chambers 1 and 2. Background *ρ* remains low for at least 1 hour after the ionizer has been switched off.

## RESULTS AND DISCUSSIONS

### Outward and inward protections

We look at four different mask designs in this study: an N95 mask, a surgical mask and two cloth masks widely distributed by the Singapore government to its population. All four masks, including the cloth masks, contain a non-woven filter layer in the middle, whose scanning electron micrographs are shown in Fig. 4. For cloth mask 2, the filter layer can be removed from the middle slot by the wearer. According to the manufacters, the surgical mask and the two cloth masks have particle filtration efficiencies (PFE) in excess of 95% for 100 nm-sized particles. However, the reported PFE values ignore the presence of gaps around the masks.

**FIG. 4.**
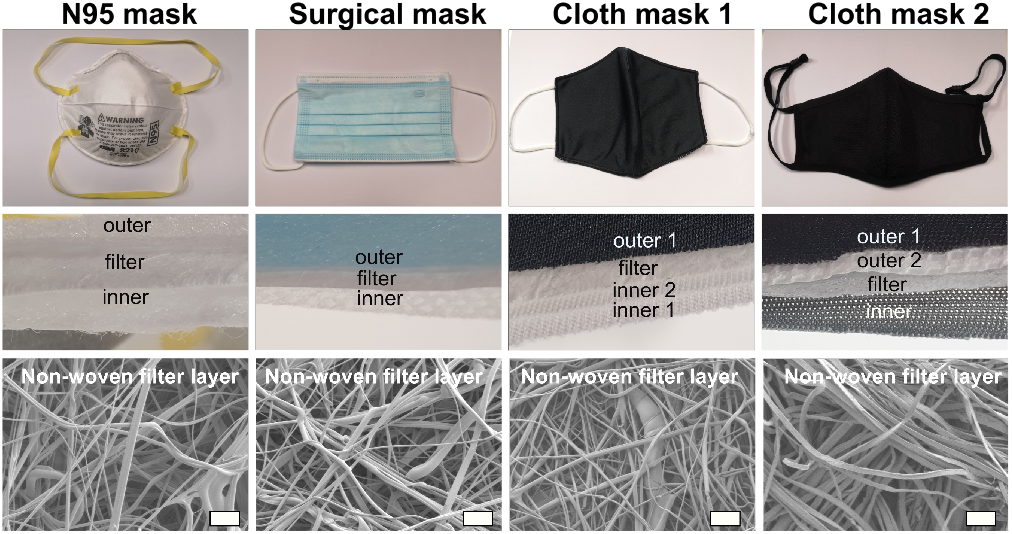
Different mask designs used in the study, consisting of three to four layers, including a non-woven filter layer. Scale bars for the scanning electron micrographs are 30 *µ*m.

A typical set of experimental results for the manikin with and without mask is summarized in Fig. 5 (dashed lines, unfilled markers and full lines, filled markers, respectively). In this case, the mask is cloth mask 2 with the filter layer removed. As discussed in the Materials and Methods section, we first generate aerosol droplets with known number densities *ρ*_1,2_ at the source chamber which we use to calculate *N*_source_(Fig. 5a–c). *ρ*_1,2_ values remain relatively unchanged during the four cycles of the simulated respiratory activities, since the volume of aerosol transferred 4*V* is much smaller than the total volume of the source chamber *V*_1,2_.

**FIG. 5.**
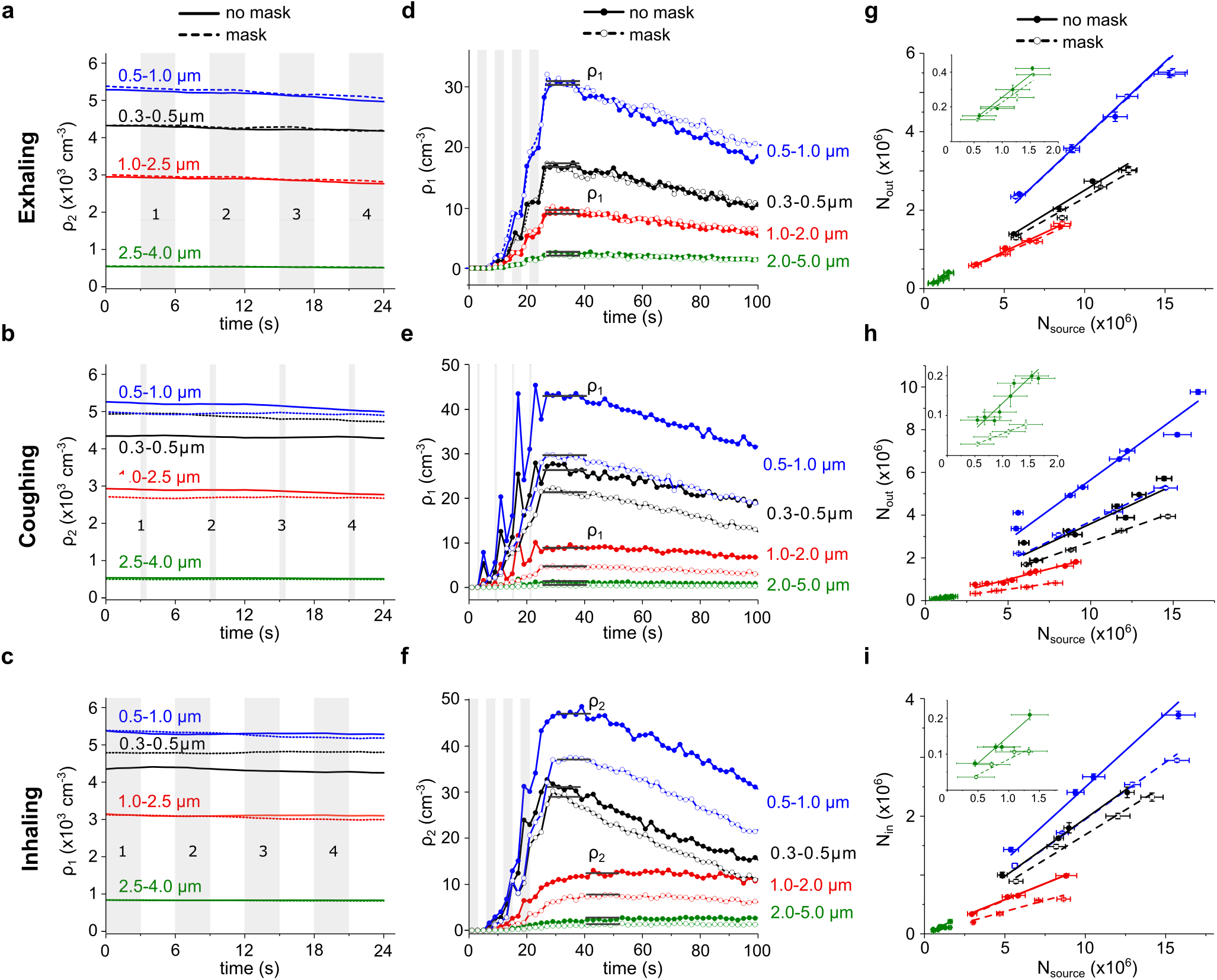
(a–c) Number density of aerosol droplets at the source: *ρ*_2_ in chamber 2 for exhaling and coughing and *ρ*_1_ in chamber 1 for inhaling. Different droplet sizes are coloured differently. The four exhalation, coughing and inhalation cycles are coloured gray and labelled 1 to 4. (d–f) Number density of aerosol droplets reaching the other chamber after simulated respiratory activities with and without mask. (g–i) Plot of total number of droplets *N*_out, in_ reaching the other chamber against the total number of droplets from the source *N*_source_. Δ*N*_out, in_ are calculated based on the standard deviation of *ρ*_2,1_(*t*) in a–c, while Δ*N*_source_ is calculated based on the uncertainty *ρ* values reported by the manufacturer for Sensirion SPS30.

With each cycle, the number densities of droplets reaching the other chamber increases until they reach a plateau (indicated by gray lines on Fig. 5d–f) whose values we use to calculate *N*_out, in_. For larger droplets *D >* 1.0 *µ*m, evaporation is unimportant and their number densities remain constant after the initial rise (red and green lines). In contrast, Ostwald ripening drives the evaporation of smaller droplets and their number densities are only constant for a few seconds before decreasing (blue and black lines) [24, 25].

For each experimental case (different respiratory activities, with and without masks), we varied the initial number of droplets from the source *N*_source_ and measured the number of droplets reaching the other chamber *N*_out, in_ (Fig. 5g–i). In our experiments, *N*_source_ ∼ 10^6^ is significantly more than the number of respiratory and aerosol droplets in a single human exhalation/cough ∼ 10^3^ [1, 26]. However, the aerosol number densities should still be considered low: for *ρ*_1,2_ ∼ 10^3^ cm^−3^, the typical spacing between neighbouring droplets *l* ∼ *ρ*^−1/3^ = 1 mm or about a thousand times the droplet diameters.

To a good approximation, the droplets do not interact with one another. Hence, we expect equations 1 and 2 to remain true and that *N*_out, in_ ∝ *N*_source_ which we verified experimentally for all the droplet sizes (Fig. 5g–i). The full and dashed lines are the lines of best fit for a linear regression with zero intercept for cases without and with mask, with lower slopes indicating better aerosol filtration. The slopes correspond to *T* for the case of no mask and (1 − *P*_out, in_)*T* with mask, from which we can calculate *P*_out, in_ using equation 4. From the various slopes in Fig. 5g–i, we can deduce that *P*_out, in_ vary widely for different respiratory activities and droplet size ranges, which will be elaborated in the rest of the manuscript.

We repeated the experiments for the four mask designs. Different masks offer different levels of protections *P*_out, in_, as evident from the different slopes of *N*_out, in_ against *N*_source_ for droplets of size range *D* = 0.75 ± 0.25 *µ*m (Fig. 6a–c). Similar experiments can be performed for the other *D* range to obtain the corresponding *P*_out, in_ (Fig. 6d–f).

**FIG. 6.**
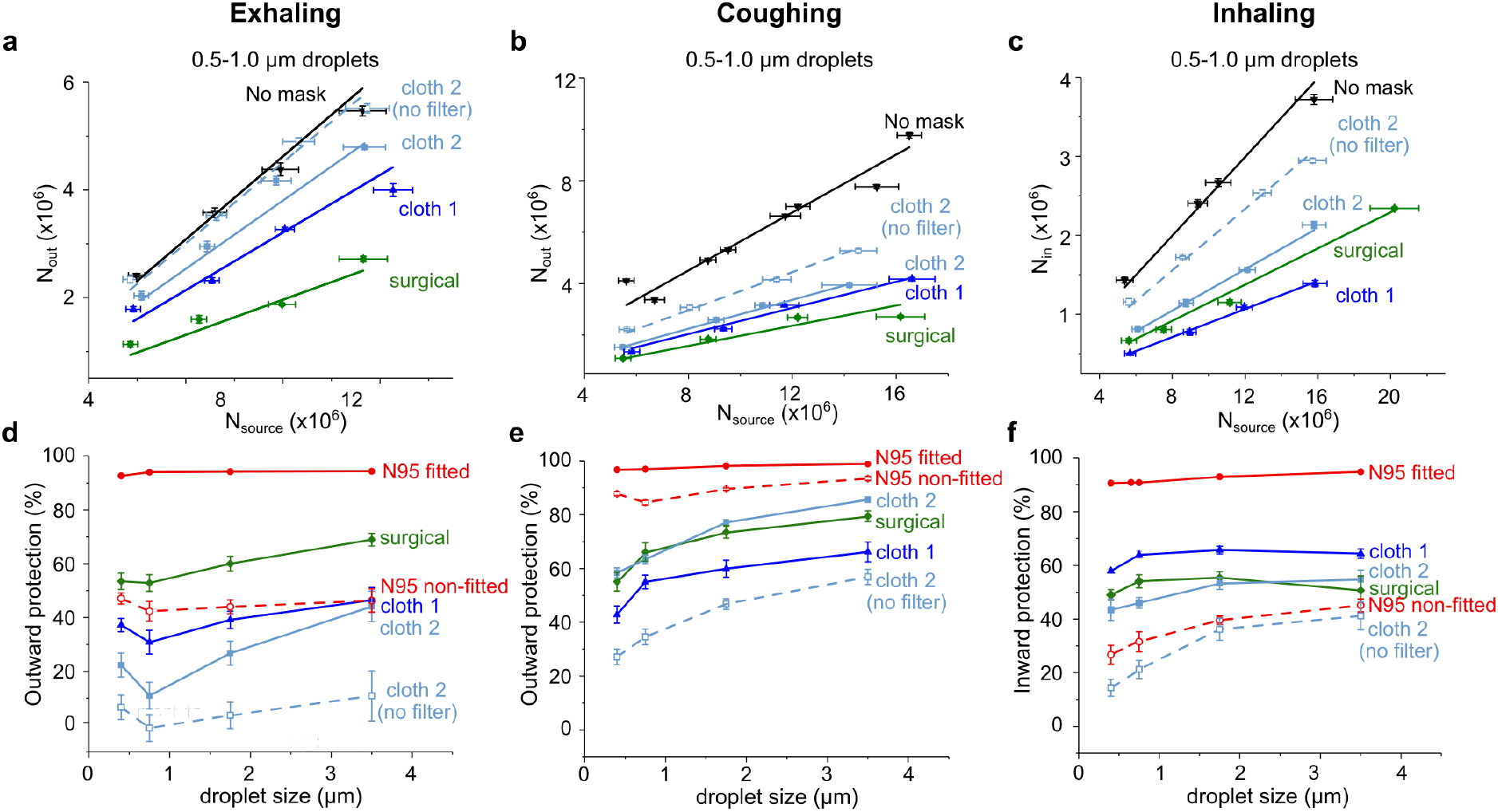
Plot of *N*_out, in_ against *N*_source_ for (a) exhaling, (b) coughing and (c) inhaling. The droplet size range here is *D*=0.75 ± 0.25 *µ*m. Lines are best-fit results from linear regression. Results for N95 masks are not shown to avoid cluttering. (d–f) Plot of *P*_out, in_ for different respiratory activities, mask designs and droplet size ranges *D* = 0.4 ± 0.1, 0.75 ± 0.25, 1.5 ± 0.5 *µ*m and 3.5 ± 1.5 *µ*m. Error bars in *P*_out, in_ are calculated using equation 5. Lines are guides to the eye.

Not surprsingly, a well-fitted N95 mask achieves *P*_out, in_ *>* 90% for all *D* range and all respiratory activities, close to the reported 95% filtration efficiency (red solid line, filled markers). In contrast, an N95 mask with sub-optimal fit (red dashed line, unfilled markers) provides little to no additional protection *P*_out_ for exhalation as compared to cloth and surgical masks (Fig. 6d); in fact, a poorly fitted N95 mask offers even less protection *P*_in_ < 40% for inhalation as compared to cloth and surgical masks (Fig. 6f). This is because more than half of the air flows through the gaps around the N95 mask unfiltered.

Cloth masks (blue lines and markers) offer similar level of protections *P*_out, in_ ≈ 50% as surgical masks (green lines and markers) as long as they contain a non-woven filter layer. Removing the filter layer significantly reduces *P*_out, in_ by a factor of 2 or more (compare light blue solid and dashed lines).

For the same flow rate *Q* = 15 L min^−1^, we found that *P*_in_ for inhalation is higher than *P*_out_ for cloth masks, consistent with results by Pan *et al*. (2021) [12]. This is likely because the negative pressure generated during inhalation draws the flexible mask closer to the face, reducing gaps. In contrast, no such effect was observed for an N95 mask due to its rigid, hard shell.

*P*_out_ for coughing are consistently highest for all mask types and *D* ranges as compared to exhalation and inhalation. This is because coughing with its higher flow rate *Q* = 140 L min^−1^ and pressure gradient forces more of the aerosol laden air through the filtration layer. As a result, a poorly fitted N95 mask is able to achieve *P*_out_ close to that of a well-fitted N95 mask (red dashed and solid lines in Fig. 6e).

Recently, the CDC reported that the wearing of a cloth mask on top of a surgical mask or double masking can help improve mask fit and reduce a wearer’s exposure by more than 90% [7]. Here, we found similar improvement in *P*_out_ for a simulated cough when wearing cloth mask 1 or 2 over a surgical mask (Fig. 7b). However, double-masking provides little to no additional protection *P*_out_ for exhaling (Fig. 7a) and less protection *P*_in_ for inhaling (Fig. 7c).

**FIG. 7.**
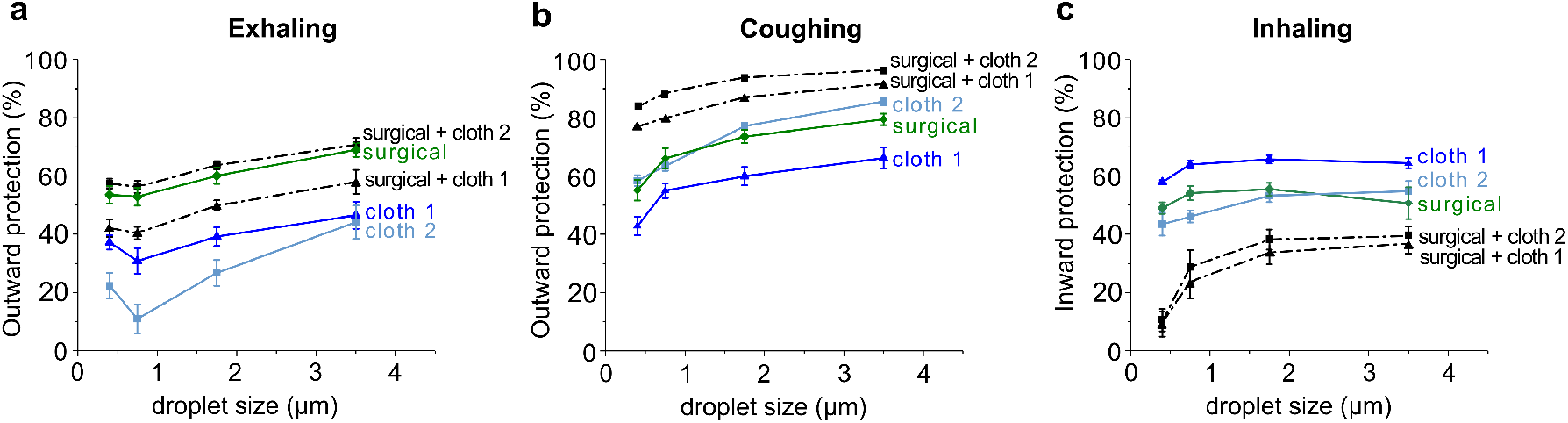
Outward protection by double masking (surgical and cloth mask as inner and outer layers, respectively) when (a) exhaling and (b) coughing. (c) Inward protection by double masking when inhaling.

Interestingly, the performance of double-masking, closely mirrors that for a poorly fitted N95 mask (compare black dashed lines in Fig. 7a–c to red dashed lines in Fig. 6d–f). This suggest that while the addition of cloth mask improves the material filtration efficiency, resulting in improved *P*_out_ for coughing, it also decreases breathability and increases material rigidity, resulting in poorer *P*_in_ for inhalation.

We can now calculate the combined outward and inward protections for different mask designs and respiratory activities

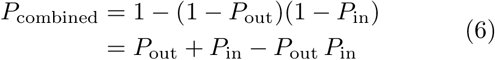

which is also the risk reduction in airborne transmissions with universal masking in the entire population.

For respiratory activities with low flow rates such as exhaling and inhaling, there is little to no benefit for the general public to wear N95 mask (Fig. 8a) or double masks (Fig. 8b). For respiratory activities with high flow rates such as coughing, there is some additional protection with poorly fitted N95 mask (Fig. 8c) and double masks (Fig. 8d), as compared to a single surgical/cloth mask. For COVID-19, it is not clear which respiratory activity (coughing/sneezing vs. exhaling) is the dominant mode of airborne transmissions.

**FIG. 8.**
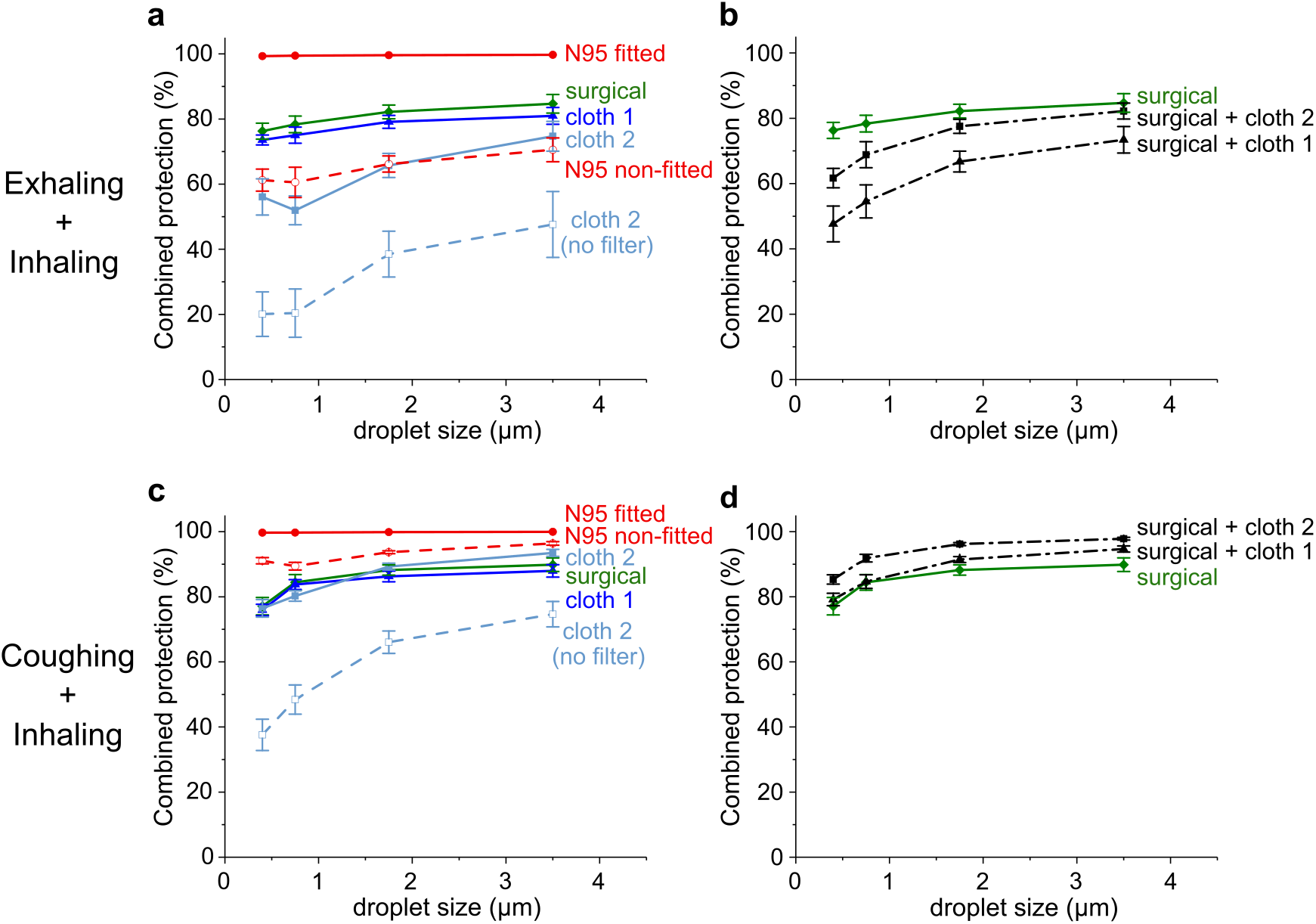
Combined outward protection when exhaling and inward protection when inhaling for (a) single masks and (b) double masks. Combined outward protection when coughing and inward protection when inhaling for (c) single masks and (d) double masks.

A well-fitted N95 masks offers the best *P*_combined_ close to 100% and should be the default choice for high-risk activities, such as healthcare workers in contact with infectious patients. For the general public, a single surgical and cloth mask offers significant protection with *P*_combined_ between 50% and 90%, as long as it contains a filter layer.

### Limitations of our study

We have only looked at the effectiveness of mask protections on a single manikin head, even though the mask fit is sensitive to head geometry. Moreover, the manikin is made of a rigid plastic material which is different from deformable human flesh. It is also likely that filtration material will degrade with regular use and washing. In future studies, we plan to address these shortcomings, for example by 3D printing manikin heads of different sizes and covering them with a soft silicone material [27].

## CONCLUSIONS

We have looked at the outward and inward protections for different mask designs. We found that while well-fitted N95 masks offer excellent protection, a poorly fitted N95 mask and double masks offer no additional or less protection as compared to a single cloth/surgical mask. For the general population, a single cloth/surgical mask with a non-woven filter layer offers significant outward and inward protections.

## Data Availability

The data that support the findings of this study are available from the corresponding author upon reasonable request.

## Authors’ contributions

D.D. conceived the research idea and supervised the research. X.Q.K. and A.S. performed the experiment, while J.Y.C. and A.S. contributed to the experimental design. P.L helped with SEM imaging. D.D. wrote up the manuscript. All authors have read and approved the manuscript.

## Acknowledgements

The authors would like to acknowledge funding from by Agency for Science, Technology and Research, Singapore (SC25/20-8R1640).

## References

[1] V. Stadnytskyi, C. E. Bax, A. Bax, and P. Anfinrud, “The airborne lifetime of small speech droplets and their potential importance in SARS-CoV-2 transmission,” Proc. Natl. Acad. Sci. U. S. A. 117, 11875–11877 (2020).

[2] G. A. Somsen, C. van Rijn, S. Kooij, R. A. Bem, and D. Bonn, “Small droplet aerosols in poorly ventilated spaces and SARS-CoV-2 transmission,” The Lancet. Respiratory Medicine, 658–659 (2020).

[3] Centers for Disease Control and Prevention, “Guidance for wearing masks help slow the spread of covid-19,” (2021).

[4] Forbes, “Germany mandates medical-grade masks,” (2021).

[5] E. Mahase, “Covid-19: What is the evidence for cloth masks?” Br. Med. J. 369 (2020), 10.1136/bmj.m1422.

[6] S. A. Grinshpun, H. Haruta, R. M. Eninger, T. Reponen, R. T. McKay, and S.-A. Lee, “Performance of an n95 filtering facepiece particulate respirator and a surgical mask during human breathing: two pathways for particle penetration,” J. Occup. Environ. Hyg. 6, 593–603 (2009).

[7] J. T. Brooks, D. H. Beezhold, J. D. Noti, J. P. Coyle, R. C. Derk, F. M. Blachere, and W. G. Lindsley, “Maximizing fit for cloth and medical procedure masks to improve performance and reduce SARS-CoV-2 transmission and exposure, 2021,” Morb. Mortal. Wkly. Rep. 70, 254 (2021).

[8] Japan Times, “Double masking has limited effect, super-computer fugaku shows,” (2021).

[9] A. Konda, A. Prakash, G. A. Moss, M. Schmoldt, G. D. Grant, and S. Guha, “Aerosol filtration efficiency of common fabrics used in respiratory cloth masks,” ACS Nano 14, 6339–6347 (2020).

[10] C. D. Zangmeister, J. G. Radney, E. P. Vicenzi, and J. L. Weaver, “Filtration efficiencies of nanoscale aerosol by cloth mask materials used to slow the spread of sarscov-2,” ACS Nano 14, 9188–9200 (2020).

[11] E. L. Kolewe, Z. Stillman, I. R. Woodward, and C. A. Fromen, “Check the gap: Facemask performance and exhaled aerosol distributions around the wearer,” PloS one 15, e0243885 (2020).

[12] J. Pan, C. Harb, W. Leng, and L. C. Marr, “Inward and outward effectiveness of cloth masks, a surgical mask, and a face shield,” Aerosol Sci. Tech., 1–17 (2021).

[13] William G Lindsley, Francoise M Blachere, Brandon F Law, Donald H Beezhold, and John D Noti, “Efficacy of face masks, neck gaiters and face shields for reducing the expulsion of simulated cough-generated aerosols,” Aerosol Science and Technology 55, 449–457 (2021).

[14] L. Bourouiba, E. Dehandschoewercker, and J. W. M. Bush, “Violent expiratory events: on coughing and sneezing,” J. Fluid Mech. 745, 537–563 (2014).

[15] Natural ventilation for infection control in health-care settings, Chap. Annex C.

[16] L. Morawska, G.R. Johnson, Z. D. Ristovski, M. Hargreaves, K. Mengersen, S. Corbett, C. Y. H. Chao, Y. Li, and D. Katoshevski, “Size distribution and sites of origin of droplets expelled from the human respiratory tract during expiratory activities,” J. Aerosol Sci. 40, 256–269 (2009).

[17] N. Wilson, S. Corbett, and E. Tovey, “Airborne transmission of covid-19,” Br. Med. J. 370 (2020).

[18] K. Tran, K. Cimon, M. Severn, C. L. Pessoa-Silva, and J. Conly, “Aerosol generating procedures and risk of transmission of acute respiratory infections to healthcare workers: a systematic review,” PloS one 7, e35797 (2012).

[19] E. P. Vejerano and L. C. Marr, “Physico-chemical characteristics of evaporating respiratory fluid droplets,” J. R. Soc. Interface 15, 20170939 (2018).

[20] D. Daniel, M. Lin, I. Luhung, T. Lui, A. Sadovoy, X. Koh, A. Sng, T. Tran, S. Schuster, X. J. Loh, O. S. Thet, and C. K. Tan, “Effective design of barrier enclosure to contain aerosol emissions from covid-19 patients,” Indoor Air (2021).

[21] L. L. Janssen, N. J. Anderson, P. E. Cassidy, R. A. Weber, and T. J. Nelson, “Interpretation of inhalation airflow measurements for respirator design and testing,” J. Int. Soc. Respir. Prot. 22, 122 (2005).

[22] J. K. Gupta, C.-H. Lin, and Q. Chen, “Flow dynamics and characterization of a cough,” Indoor Air 19, 517–525 (2009).

[23] K. K. Lee, S. Matos, K. Ward, G. F. Rafferty, J. Moxham, D. H. Evans, and S. S. Birring, “Sound: a non-invasive measure of cough intensity,” BMJ Open Respir. 4 (2017).

[24] P. W. Voorhees, “Ostwald ripening of two-phase mixtures,” Annu. Rev. Mater. Sci. 22, 197–215 (1992).

[25] T. H. Tsang and J. R. Brock, “On ostwald ripening,” Aerosol Sci. Tech. 3, 283–292 (1984).

[26] G.R. Johnson, L. Morawska, Z. D. Ristovski, M. Hargreaves, K. Mengersen, C.Y.H. Chao, M.P. Wan, Y. Li, X. Xie, D. Katoshevski, and S. Corbett, “Modality of human expired aerosol size distributions,” J. Aerosol Sci. 42, 839–851 (2011).

[27] M. S. Bergman, Z. Zhuang, D. Hanson, B. K. Heimbuch, M. J. McDonald, A. J. Palmiero, R. E. Shaffer, D. Harnish, M. Husband, and J. D. Wander, “Development of an advanced respirator fit-test headform,” J. Occup. Environ. Hyg. 11, 117–125 (2014).

